# Evaluation of DBS computational modeling methodologies using in-vivo electrophysiology in Parkinson’s disease

**DOI:** 10.1101/2025.05.05.25326314

**Authors:** Seyyed Bahram Borgheai, Bryan Howell, Faical Isbaine, Angela M Noecker, Enrico Opri, Benjamin B Risk, Cameron C McIntyre, Svjetlana Miocinovic

## Abstract

Deep brain stimulation (DBS) is an effective therapy for Parkinson’s disease (PD) and other neuropsychiatric disorders, but its outcomes vary due to differences in patient selection, electrode placement, and programming. Optimizing DBS parameter settings requires postoperative adjustments through a trial-and-error process, which is complex and time-consuming. As such, researchers have been developing patient-specific computational models to help guide DBS programming. Despite growing interest in image-guided DBS technology, and recent adoption into clinical practice, the direct validation of the prediction accuracy remains limited. The objective of this study was to establish a comparative framework for validating the accuracy of various DBS computational modeling methodologies in predicting the activation of clinically relevant pathways using in vivo measurements from PD patients undergoing subthalamic (STN) DBS surgery. Our prior work assessed the accuracy of driving force (DF) models in native space by predicting activation of the corticospinal/bulbar tract (CSBT) and cortico-subthalamic hyperdirect pathway (HDP) using very short-(<2 ms) and short-latency (2–4 ms) cortical evoked potentials (cEPs). In this study, we extended our previous work by comparing the accuracy of five computational modeling variations for predicting the activation of HDP and CSBT based on three key factors: modeling method (DF vs. Volume of Tissue Activated [VTA]), imaging space (native vs. normative), and anatomical representation (pathway vs. volume). The model performances were quantified using the coefficient of determination (R^2^) between the cEP amplitudes and percent pathway activation or percent volume (structure) overlap. We compared model accuracy for 11 PD patients. The DF-Native-Pathway model was the most accurate method for quantitatively predicting experimental subcortical pathway activations. Additionally, our analysis showed that using normative brain space, instead of native (i.e., patient-specific) space, significantly diminished the accuracy of model predictions. Although the DF and VTA modeling methods exhibited comparable accuracy for the hyperdirect pathway, they diverged significantly in their predictions for the corticospinal tract. In conclusion, we believe that the choice of methodology should depend on the specific application and the required level of precision in clinical, surgical, or research settings. These findings offer valuable guidance for developing more accurate models, facilitating reliable DBS outcome predictions, and advancing both clinical practice and research.

## Introduction

Deep brain stimulation (DBS) is an effective therapy for Parkinson’s disease (PD) and is being explored for various neuropsychiatric disorders. However, DBS outcomes vary due to differences in patient selection, electrode placement, and device programming. Its efficacy largely depends on postoperative programming to optimize activation of target structures while minimizing side effects. Currently, this involves an iterative trial-and-error process across a vast parameter space, making programming complex and time-consuming (Krack et al., 2002). This is made more challenging by technological advances such as segmented leads which have further expanded the parameter space (ten Brinke et al., 2018). Computational models of DBS are of increased interest to address these limitations through in-silico approaches, reducing the need for continuous patient interaction. These 3D, image-based, biophysical models visualize neuronal activation patterns around electrodes and connected regions (Janson & Butson, 2018). Pilot studies suggest they can reduce programming time (Pavese et al., 2020; Pourfar et al., 2015) and improve outcomes (Frankemolle et al., 2010). Modeling DBS effects in 3D is also valuable for understanding neural pathways and mechanisms involved in therapeutic effects and side effects (Mädler & Coenen, 2012; Nguyen et al., 2019; Riva-Posse et al., 2014; Schaper et al., 2020; Sweet et al., 2014).

Despite growing interest in image-guided DBS programming and recent adoption into clinical practice, the direct validation of model prediction accuracy remains limited. For instance, studies correlating model-defined ‘sweet spots’ with clinical outcomes have shown statistical significance but low correlation (Dembek et al., 2019; Elias et al., 2021; Horn et al., 2017; Treu et al., 2020) and the models are generally poor at predicting stimulation-induced side effects (Béreau et al., 2020). In addition, wide-ranging variability in the technical modeling methodology translates into a lack of standardization that has hindered validation (Calabrese, 2016; Dembek et al., 2022)(Calabrese, 2016). However, standardizing DBS modeling to more coherently explain clinical effects requires revisiting fundamental assumptions. Current models may fall short due to their ‘black box’ approach, attempting to directly link electric field values to clinical outcomes. Since DBS outcomes are closely tied to neural activations, particularly of myelinated axons (McIntyre et al., 2004; Nowak & Bullier, 1998a, 1998b), first, models must accurately estimate activations of relevant structures and pathways. So, to make the use of DBS models justifiable in clinical research and successful in clinical practice, a rigorous and systematic optimization and validation is necessary to demonstrate that DBS models accurately estimate activation of related structures and pathways.

In this study, we focus on three key sources of variability in DBS modeling: imaging space, biophysical modeling method, and anatomical representation. Patient-specific pipelines utilize native imaging space to define individual anatomy, electrode placement, and pathway trajectories, enabling tailored neural activation models (Anderson et al., 2018; Gunalan et al., 2017). Alternatively, co-registering patient imaging into a normative space enables the use of standardized atlases, facilitating group-level analyses like probabilistic “sweet spot” mapping for therapeutic efficacy (Akram et al., 2017; Butson et al., 2011; Dembek et al., 2019; Horn et al., 2017). Biophysical modeling of neuronal activation around stimulating contact also differs in complexity. Simplified volume of tissue activation (VTA) models estimate activation using uniformly distributed axons around the electrode (Butson et al., 2006b) and fixed electric field thresholds as proxies for axonal activation in symmetric/isotropic fields (Åström et al., 2015a; Duffley et al., 2019; Horn et al., 2019), therefore neglecting patient-specific information and antisotropic properties of the surrounding tissues (Duffley et al., 2019; Horn et al., 2019). Driving force (DF) models, in contrast, leverage electric field gradients and more realistic axonal trajectories derived from tractography or pathway atlases (Gunalan et al., 2017; Howell, Choi, et al., 2019). Anatomical representation is another factor distinguishing DBS modeling. VTA models traditionally use structure-based atlases, assuming activation overlaps with target regions, while DF models and more recent VTA approaches incorporate pathway (connectomic) atlases to account for individual axons passing near the electrode (Howell et al., 2021; Rajamani et al., 2024). Despite these advancements, no comparative studies have definitively determined which imaging space, modeling method, or anatomical representation provides superior accuracy or clinical utility.

The objective of this study is to establish a comparative framework for validating the accuracy of various DBS computational modeling methodologies in predicting the activation of clinically relevant pathways using in vivo measurements from PD patients undergoing subthalamic (STN) DBS surgery. We employed cortical evoked potentials (cEPs) as an objective gold standard of pathway activation, as previous studies have shown that the latencies and amplitudes of cEPs evoked by DBS can reveal which pathways near the STN are activated (Borgheai et al., 2025; Jorge et al., 2022; Miocinovic et al., 2018). Our prior work assessed the accuracy of DF models in native space in predicting activation of the corticospinal/bulbar tract (CSBT) and cortico-subthalamic hyperdirect pathway (HDP) using very short-(<2 ms) and short-latency (2–4 ms) cEPs, as their respective experimental measures (Howell et al., 2021). In this study, we extended that work by comparing different DBS modeling methodologies, identifying the impact of key factors, namely imaging space, modeling method, and anatomical representation. Our findings aim to guide the development of more accurate models, ultimately guiding their appropriate use in both DBS clinical practice and research.

## Methods

### Patient selection

Patients diagnosed with idiopathic PD and scheduled for awake STN DBS surgery at two major academic centers participated in the study (Table 1). Prior to surgery, written informed consent was obtained in accordance with protocols approved by the Institutional Review Boards. Patients were informed that a temporary subdural electrocorticography (ECoG) recording electrode would be inserted exclusively for research purposes during the procedure.

**Table 1.** Patient demographics and experimental setup.

| Patient code | Age/Sex | ECoG laterality at M1 (mm) | Number of monopolar DBS settings modeled | Number of bipolar DBS settings modeled | DBS lead | Lead type | DBS lead bottom contact MCP coordinates (mm) |  |  |
| --- | --- | --- | --- | --- | --- | --- | --- | --- | --- |
| P01 | 6X/M | 34.0 | 14 | 22 | MDT 3389 | Standard | 12.1 | -4.9 | -3.5 |
| P02 | 5X/M | 40.9 | 14 | 28 | MDT 3389 | Standard | 8.8 | -1.1 | -4.6 |
| P03 | 5X/M | 24.3 | 14 | 25 | MDT 3389 | Standard | 10.8 | -3.9 | -2.8 |
| P04 | 4X/M | 18.9 | 33 | 9 | ABT 6172 | Steerable | 9.8 | -4.9 | -5.9 |
| P05 | 5X/M | 28.0 | 39 | 10 | BSC 2202 | Steerable | 10.7 | -2.5 | -6.6 |
| P06 | 6X/M | 26.5 | 34 | 6 | ABT 6172 | Steerable | -12.7 | -2.3 | -4.2 |
| P07 | 6X/M | 35.0 | 0 | 18 | MDT 3389 | Standard | 13.1 | -5.4 | -6.1 |
| P08 | 6X/M | 44.0 | 14 | 20 | MDT 3389 | Standard | 13.4 | -1.3 | -4.1 |
| P09 | 6X/M | 41.5 | 0 | 18 | MDT 3387 | Standard | 12.7 | -2.4 | -4.0 |
| P10 | 6X/M | 27.5 | 9 | 18 | BSC 2201 | Standard | 9.3 | -4.8 | -7.3 |
| P11 | 6X/M | 22.6 | 0 | 15 | MDT 3387 | Standard | 10.9 | -1.6 | -4.6 |

M1 = primary motor cortex; MCP = mid-commissural point; ABT= Abbott; BSC = Boston Scientific; MDT= Medtronic.

### Experimental setup and signal acquisition

In this study, we used the same experimental dataset reported in our previous work (Howell et al., 2021). Briefly, cortical evoked potentials (cEP) were recorded with a subdural ECoG strip (28 or 6 contacts; Ad-Tech) placed on the surface of the brain via the same burr hole used for DBS implantation (Miocinovic et al., 2018; Panov et al., 2017) (Figure 1-A). The intended target location for the center of the strip was the arm area of M1, ∼3 cm from the midline and slightly medial to the hand knob. Recordings were performed at least 12 hours after stopping all anti-parkinsonian medications and at least 30 minutes after stopping propofol sedation. ECoG electrode location was determined using intraoperative CT registered to the preoperative MRI in surgical planning software (Framelink 5.1, Medtronic). Contact pairs overlying the precentral gyrus (M1) were used for further analysis (Figure 1-A). ECoG potentials were recorded using either the Neuro Omega (Alpha Omega Engineering) at a sampling rate of 22 kHz or TDT PZ5 (Tucker Davis Technologies; P09 and P11) acquisition systems at a sampling rate of 24,414 Hz. Ear electrode (P01-P06) or an ipsilateral scalp needle (P07-P11) was used as the reference with the corresponding contralateral electrode as the ground.

**Figure 1.**
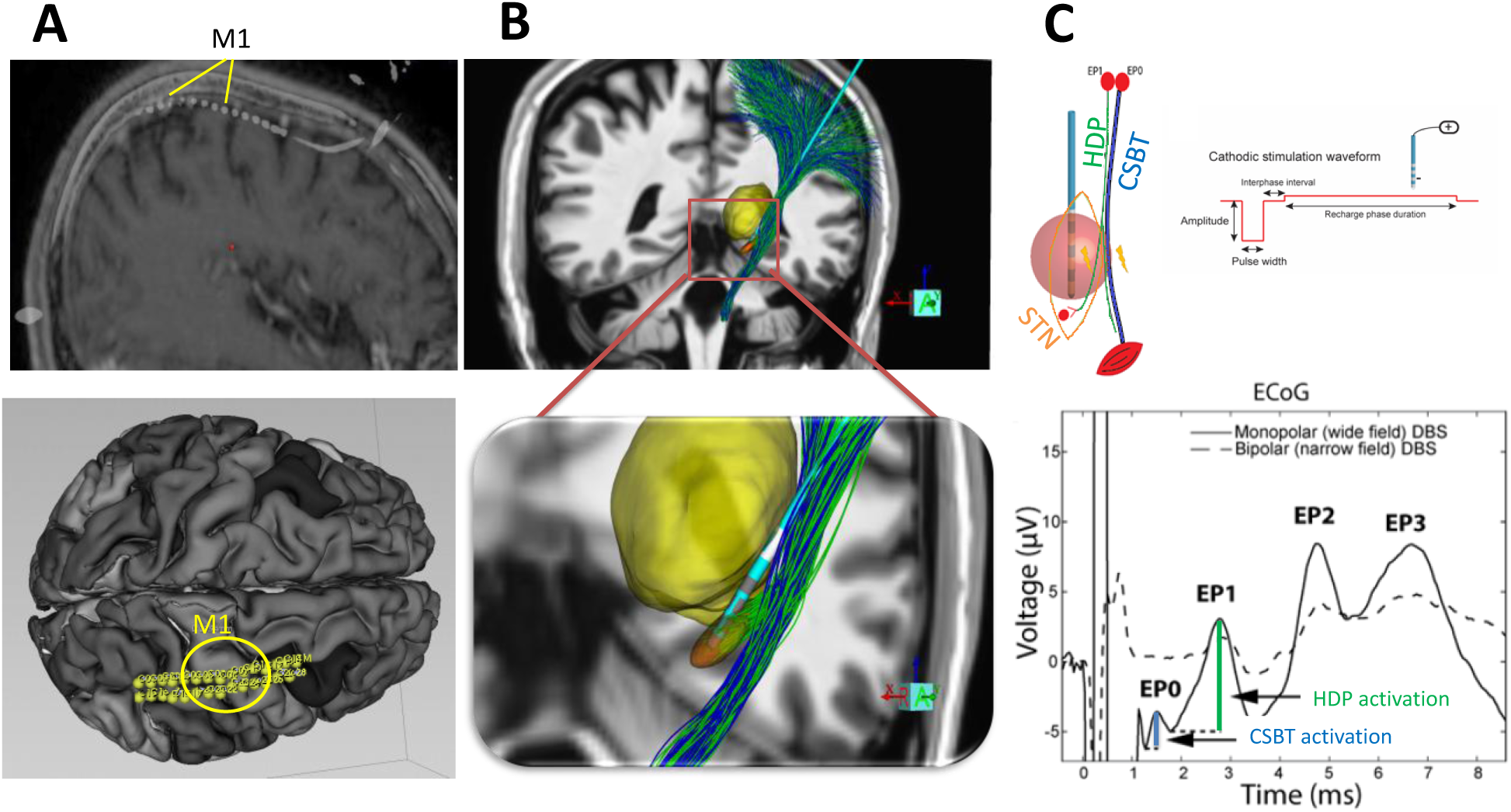
Experimental Setup and Target Pathways. A) Top: Sagittal view of a temporary 28-contact subdural electrocorticography (ECoG) strip placed over the M1 region. Bottom: Top-down view of the ECoG placement visualized by Slicer 5.2. B) Modeling of the DBS electrode implanted in the subthalamic nucleus (STN) (orange), shown in relation to the thalamus (yellow), the hyperdirect pathway (HDP, green), and the cortico-spinal bulbar tract (CSBT, blue), visualized by StimVision. C) Top: Schematic representation of HDP and CSBT antidromic activation in response to STN stimulation. Bottom: Example of a cortical evoked potential (cEP) recorded from one of the ECoG strip contacts in response to STN stimulation. The first and second peak amplitudes, referred to as very short-latency evoked potential (EP0) and short-latency evoked potential (EP1) (Miocinovic et al., 2018), are used here as measures of HDP and CSBT activation. STN= subthalamic nucleus; HDP = hyperdirect pathway; and CSBT = corticospinal/bulbar tract.

### Experimental Stimulation and Recordings

We experimentally measured antidromic activation of the hyperdirect pathway (HDP, shown in green) and the corticospinal-bulbar tract (CSBT, shown in blue) generated by STN DBS (Figure 1B and C, top). Activation of these pathways was quantified using very short-latency (<2 ms) and short-latency (2–4 ms) cortical evoked potential amplitudes, denoted as EP0 and EP1, respectively (Figure 1C, bottom). The cEP amplitudes have been previously demonstrated to indicate the degree of HDP and CSBT activation (Borgheai et al., 2025; Miocinovic et al., 2018).

The number of stimulation settings tested varied across patients (range 15-49) due to logistical constraints and available intraoperative time (Table 1). We used the same datasets and stimulation settings as in our previous study (Howell et al., 2021). However, only cathodic configurations were included in this study (Figure 1C, top) because the software we used for VTA-based modeling (Lead DBS v3.1) could not model anodic configurations. All stimulation waveforms (except for three settings) were asymmetric biphasic, with a 70 µs inter-phase interval (Figure 1C, top), and were applied for 12 seconds at 10 Hz resulting in ∼120 pulses.

### Neuroimaging and Lead Localization

Preoperative T1-weighted (T1w) MRI scans were obtained using 1.5T or 3T scanners, as detailed in (Howell et al., 2021). For DF-based models, the FMRIB Software Library (FSL) (Smith et al., 2004) was used for neuroimaging analysis. T1w images were corrected for RF/B1 inhomogeneities with FMRIB’s Automated Segmentation Tool (FAST) (Zhang et al., 2001) and underwent skull stripping using FSL’s Brain Extraction Tool (BET) (Nakamura et al., 2005). Pre- and post-operative MRIs registration was performed using FSL’s FLIRT (Jenkinson et al., 2002; Jenkinson & Smith, 2001). In cases where FLIRT resulted in noticeable distortions in the ventricles or major anatomical structures—such as the cerebellum, corpus callosum, or large gray matter regions—we instead used FNIRT (Andersson et al., 2007). Images were co-registered with a rigid-body affine transformation and Normalized Mutual Information as the cost function, unless registration was poor, in which case a twelve-parameter affine transformation was applied (for P01, P03, and P06).

For VTA-based models, the postoperative T1 were co-registered and resliced to the upsampled T1 using SPM 12 and Advanced Normalization Tools (ANTs) through built-in modules in Lead-DBS V3.1(Neudorfer et al., 2023). Co-registration results were then visually inspected using built-in tools. To compensate for deformation of the brain during surgery, we also applied the brain-shift correction method embedded in Lead-DBS (Horn et al., 2019; Neudorfer et al., 2023).

The brain atlases used to estimate anatomical volumes or pathways within the imaging data were processed using a pipeline detailed in the Anatomical Representation section below. Postoperative T1-weighted images were used for lead localization. This choice was driven by the availability of postoperative MRI images across all patients (except P10, where postoperative CT was used instead).

For DF-based models, lead localization was performed using a custom MATLAB script with thresholding and orthogonal distance regression (ODR) (Howell et al., 2021). To make sure that DBS contact localization was consistent across different computational models, identified contact locations were marked (‘burned’) in the postoperative T1w images using 3D Slicer v5.2.1 and imported into Lead-DBS for VTA-based modeling. Using the manual lead localization tool in Lead-DBS, we then aligned the electrode contacts to the marked points marked in the postoperative T1w images. This approach ensured consistency in electrode positions between the DF-based and VTA-based modeling frameworks.

### Computational Modeling Variations

We implemented 5 model variations based on combination of 3 key factors: imaging space, modeling method, and anatomical representation (Figure 2). The modeling method was either DF or VTA. The imaging space was either native (a.k.a. patient-specific) or normative (atlas-based). Neural activations in VTA-based models were calculated using either volumetric structures (overlap of anatomical structure with the VTA) or pathway streamlines (number of neural streamlines that pass through the VTA). For DF-models, pathways were the only viable anatomical representation due to their inherent focus on axonal activation predictions. In native space, we exclusively employed volume atlas-based VTA modeling. This limitation arose because pathway-based VTA modeling in native space is neither supported by the tools used in this work nor a common practice in the field. We therefore created 5 model variations: 1) DF-Native-Pathway, 2) DF-Normative-Pathway, 3) VTA-Native-Volume, 4) VTA-Normative-Volume, and 5) VTA-Normative-Pathway.

**Figure 2.**
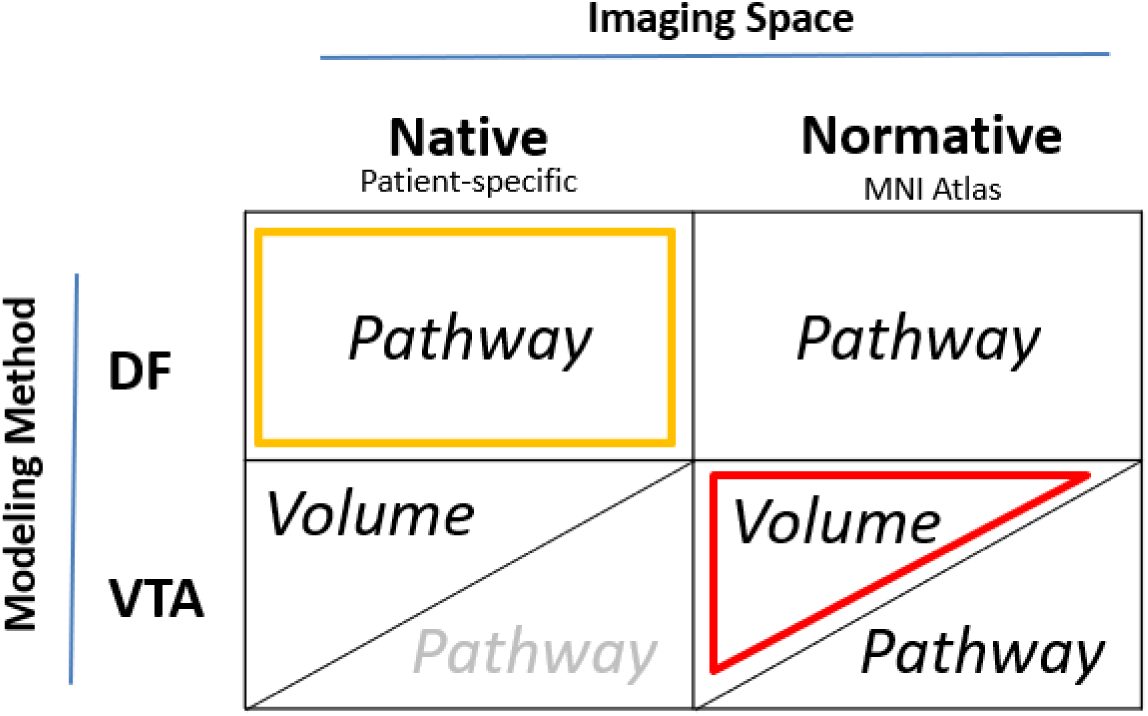
Computational model variations based on three factors: Modeling methods (VTA or DF), Imaging space (Native or Normative), and Anatomical representation (Volume or Pathway). DF by definition only models pathway activations, so there is no DF with volume-based atlases. The VTA-Native pathway modeling (pale gray) is not currently adaptable to the Lead-DBS software pipeline. The highlighted models (in yellow and red) are the most commonly used in published modeling studies .

#### Modeling Methods

##### Volume of Tissue Activation (VTA)

The VTA is a binary volume surrounding the stimulating contact, predicted to activate axons and elicit action potentials within it (Butson & McIntyre, 2005). We used Lead DBS v3.1 to construct VTAs around the stimulating leads. That software employs the SimBio/FieldTrip pipeline (Vorwerk et al., 2018) with the finite element method (FEM) to solve the Laplace equation in discretized native patient space (Fig. 3, Top-Left). Using default parameters (and ‘electrode removed’ option), we generated the VTA for each stimulation setting in each patient. The typical threshold of 0.2 V/mm was applied to the E-Field magnitude to create the VTA (Åström et al., 2015b; Horn et al., 2019; Neudorfer et al., 2023). Since the software, in its default setting, does not account for stimulation frequency or pulse width differences, changes in these parameters produced the same VTA. The VTAs were then used to calculate the percentage overlap with atlas structure volume or to calculate the number of pathway streamlines passing through the VTA as a measure of neural activation (Fig. 3, Top-Right).

**Figure 3.**
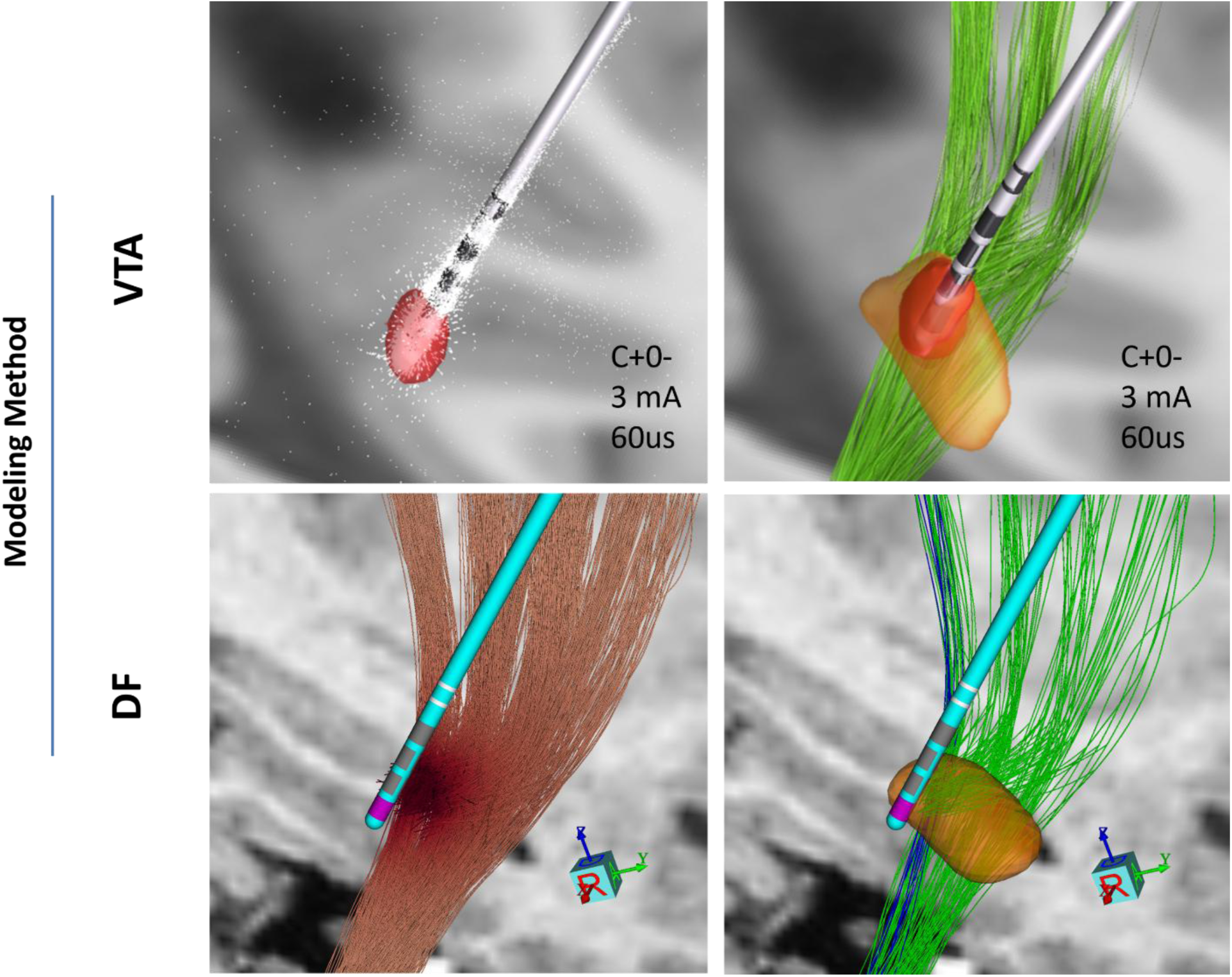
Example of different modeling methods for one patient (P06): Top images are generated in Lead DBS V3.1 and bottom ones in StimVision. Top-Left: Volume of Tissue Activated (VTA) estimate in red in response to monopolar stimulation at ventral-most contact (grey). White arrows represent electric field lines; Top-Right: VTA overlap with STN structure (orange) and HDP (green); Bottom-Left: Driving Force (DF) estimate of voltage distribution gradients along the axonal streamlines; Bottom-Right: Estimate of activated pathway streamlines for HDP (green) and CSBT (blue) in response to the same stimulation setting as above.

##### Driving Force (DF)

DF predictors were constructed as described in our prior study (Howell et al., 2021). Electric field models were built in COMSOL (v5.1). The volume conductors for DBS leads were modeled with Dirichlet and mixed boundary conditions for active and inactive contacts, respectively. Tissue models for standard and steerable leads were discretized in 2D and 3D, respectively, for efficiency. Electric potentials were calculated by solving Laplace’s equation using the Finite Element Method (FEM). Temporal variations were approximated with a waveform derived from an equivalent circuit model (Noecker et al., 2021; Vorwerk et al., 2018). Axonal stimulation thresholds were estimated using a predictive algorithm based on the driving force at the nodes of Ranvier within the CSBT and HDP (Howell, Gunalan, et al., 2019).

#### Imaging space

Each modeling method was implemented in either native (patient-specific) or normative (atlas-based) space (Fig. 4).

**Figure 4.**
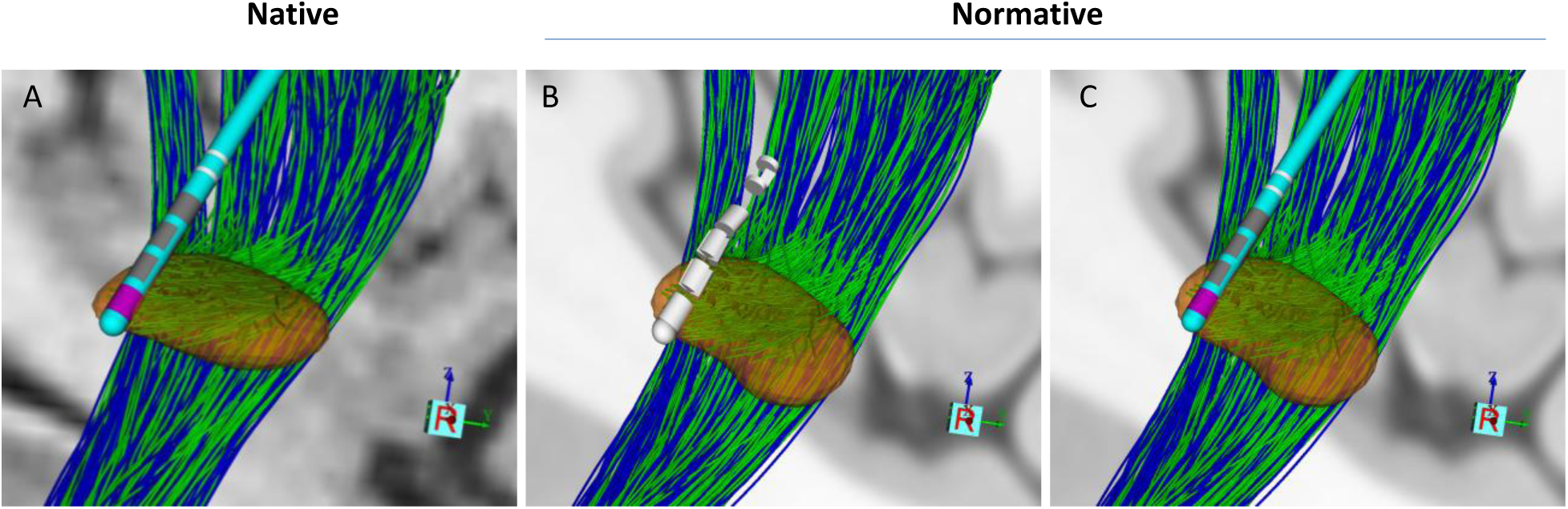
Example of modeling in different imaging spaces for one patient (P06). The normative space used here is MNI2009b. All images are generated in StimVision. The lead type is ABT6172, and STN, HDP, and CSBT are shown in orange, green, and blue, respectively. Transformation into normative space introduces visually subtle warping of the anatomical structures and the lead shaft. A) Illustration of the STN, lead, and pathways in native space, B) Warped image in normative space before fitting a lead shaft to a straight line between the contacts and tip. C) Warped image in normative space after fitting a lead shaft to a straight line between the contacts and tip.

##### Native

For DF native models, the standard CIT168 (Pauli et al., 2018) brain atlas was registered to the patient’s preoperative T1w space using FSL’s nonlinear registration tool, FNIRT (Andersson et al., 2007), with all registrations visually confirmed for quality. Additionally, CSBT and HDP were modeled using an anatomical pathway atlas of the subthalamic region (Fig. 3) defined in the CIT168 space (Petersen et al., 2019) individualized by nonlinearly warping the atlas data from CIT168 space to the patient’s preoperative T1w (native) space.

For VTA native models, the VTAs were first calculated in native space using an adaptation of the SimBio/FieldTrip pipeline (Vorwerk et al., 2018) in Lead DBS v3.1 which is the static formulation of the Laplace equation solved in native patient space.

##### Normative

For normative space, to provide a common ground between DF and VTA methods, all the imaging and atlases were warped into ICBM 2009b Nonlinear Asymmetric space known as Montreal Neurological Institute (MNI) space (Fonov et al., 2009). This version of normative space was the updated version of MNI space in 0.5 × 0.5 × 0.5 mm^3^ resolution constructed through nonlinear co-registration of 152 acquisitions (Fonov et al., 2011; Horn et al., 2019).

#### Anatomical representation and activation measures

Two anatomical representations were used when estimating the effects of DBS: Pathway or Volume.

##### Pathway

The same pathway atlas was used for VTA and DF models previously described by Petersen et al. (2019). For DF models, each pathway fiber whose activation threshold was below the stimulation amplitude was counted as activated by that stimulation setting. The percentage of fibers activated for HDP and CSBT was used as a metric of activation for each pathway (Fig. 3-Bottom-Right). We set fiber diameters as 12 mm for CSBT and 4mm for HDP based on our prior work which showed this was the optimal diameter to accurately model experimental activations (Howell et al., 2021).

For VTA models, we calculated the percentage of fibers passing through VTAs using custom MATLAB code. We first converted the VTA NIfTI file format produced in Lead-DBS into a graphical object with surfaces and vertices using *isosurface* MATLAB function. Then for each point defining a fiber trajectory, we checked whether it resided inside the VTA object using *inpolyhedron* function. If at least one point was located inside the VTA, that fiber was considered to be activated (Fig. 5-Bottom).

**Figure 5.**
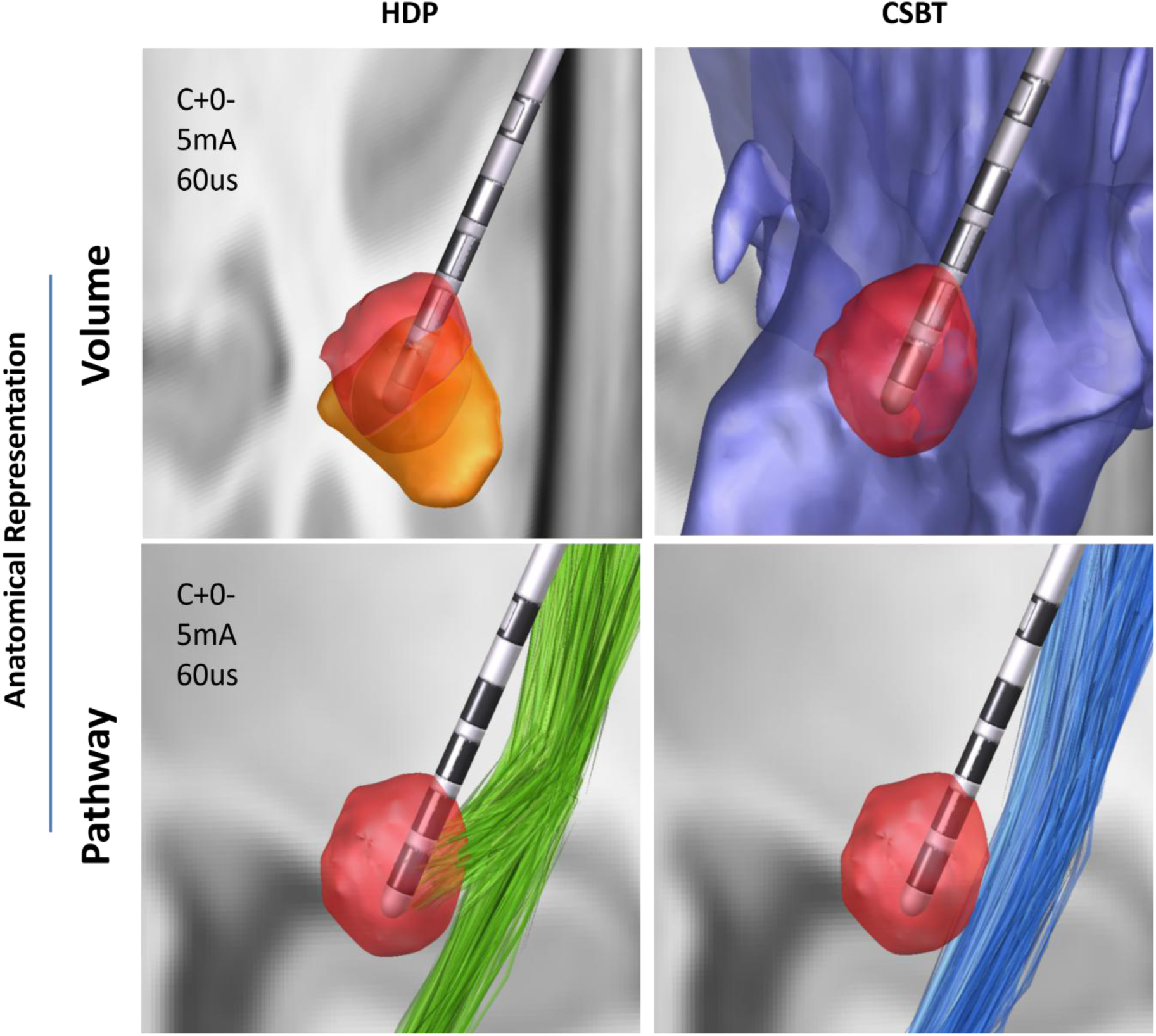
Example of different anatomical representations for one patient (P06). Top row illustrates the volume representation using Distal atlas showing overlaps of a VTA with STN (orange-Left) and IC (purple-Right). Bottom row illustrates the intersection of a VTA in response to the same stimulation with HDP (in green-Left) and CSBT (in blue-Right).

##### Volume

The overlap of VTAs with internal capsule (IC) and STN structures in the DISTAL atlas (Ewert et al., 2018) were calculated as activation metrics for CSBT and HDP, respectively, in both normative (MNI) and native spaces (Fig. 5-Top). In the normative space, we calculated the percentage of overlap volume with respect to the desired structure volume (i.e. IC and STN), using the voxel-based values reported in Lead-DBS (Horn et al., 2019; Neudorfer et al., 2023). For the overlaps in the native space, we developed a custom MATLAB code where we calculated the percentages of the structure (IC or STN) vertices located inside VTA objects in response to each DBS setting. The rationale behind this approach was that since the voxel size was the same for the VTA and structures, the ratio of overlapped vertices should be similar to the ratio of overlapped volumes (voxels). To ensure that our new overlap percentage calculation method in native space was consistent with the method used in Lead-DBS (available only for normative models), we recalculated the overlap volume percentage in normative space using the custom code and correlated with Lead-DBS overlap values. Across 5 patients (P01-05), the average correlation coefficients between the two methods were 0.990 ±0.008 and 0.916 ±0.117 for STN and IC structures, respectively.

### Statistical analysis

The model performance was quantified using the coefficient of determination (R²), i.e., the square of Pearson correlation coefficient (R), between the experimental measures of HDP and CSBT pathway activation (i.e., cEP amplitudes) and the corresponding estimates of pathway activation produced by each computational model. The predictive accuracy of all modeling variations was assessed relative to the DF-Native-Pathway described in our previous work (Howell et al., 2021) using Wilcoxon signed-rank test to compare the corresponding R² values across all patients for the HDP and CSBT pathways, with a significance level of α = 0.05. We first compared the two most common modeling strategies in the DBS literature: DF-Native-Pathway and VTA-Normative-Volume. Next, to investigate the influence of different modeling factors (imaging space, modeling method, and anatomical representation), we performed comparisons using the Wilcoxon signed rank on R² values of two models that were identical in all aspects except for the factor under investigation. Specifically, to evaluate the impact of imaging space, we compared DF-Native-Pathway with DF-Normative-Pathway and VTA-Native-Volume with VTA-Normative-Volume for each pathway. The null hypothesis for these comparisons was that there is no difference between the R² values of native and normative paired-methods.

For the CSBT pathway, in addition to R², we compared different models using F-score values, based on presence or absence of activation, to account for the imbalance in the number of true negatives in our data. This imbalance resulted from the presence of numerous zero values in either the EP0 amplitudes or the model’s estimated values. In F-score calculation, we excluded three patients whose EP0 was equal to zero for all stimulation settings.

## Results

We compared performance of different DBS computational models in predicting HDP and CSBT activations using cortical evoked potentials from 11 PD patients as the experimental activation metric. In each patient, for each stimulation setting (360 total (Table 1)), we estimated the activation of HDP and CSBT pathways using five model variations: DF-Native-Pathway, DF-Normative-Pathway, VTA-Normative-Pathway, VTA-Normative-Volume, and VTA-Native-Volume. The average number of stimulation settings tested and modeled per patient was 32 (range: 15–49) (Table 1). The model performance was evaluated using the coefficient of determination (R²) to quantify the relationship between measured cEP amplitudes and model predictions (either percent pathway activation or activation volume overlap), for two pathways of interest (HDP and CSBT).

First, we compared two most commonly used model variations, DF-Native-Pathway and VTA-Normative-Volume (Fig. 6). The median model performance was higher using the DF method, for both pathways. For HDP, the *R*^2^ was 0.73 (IQR: 0.65-0.81) for DF and 0.40 (IQR: 0.33-0.57) for VTA. This difference was statistically significant (p-value = 0.019). For CSBT, the *R*^2^ median was 0.59 (IQR: 0.47-0.70) for DF and 0.14 (IQR: 0.02-0.30) for VTA. This difference was statistically significant (p-value = 0.016) model. The VTA model performance was particularly poor for estimating the activation of CSBT pathway (modeled as overlap between the VTA and internal capsule structure). For CSBT, F-score median was 0.77 (IQR: 0.35-0.86) for DF and statistically comparable to 0.55 (IQR: 0.42-0.80) for VTA (p-value= 0.945) (Fig. S1). Overall, DF-Native-Pathway model performed better than all other models across all patients (Table S1 and S2).

**Figure 6.**
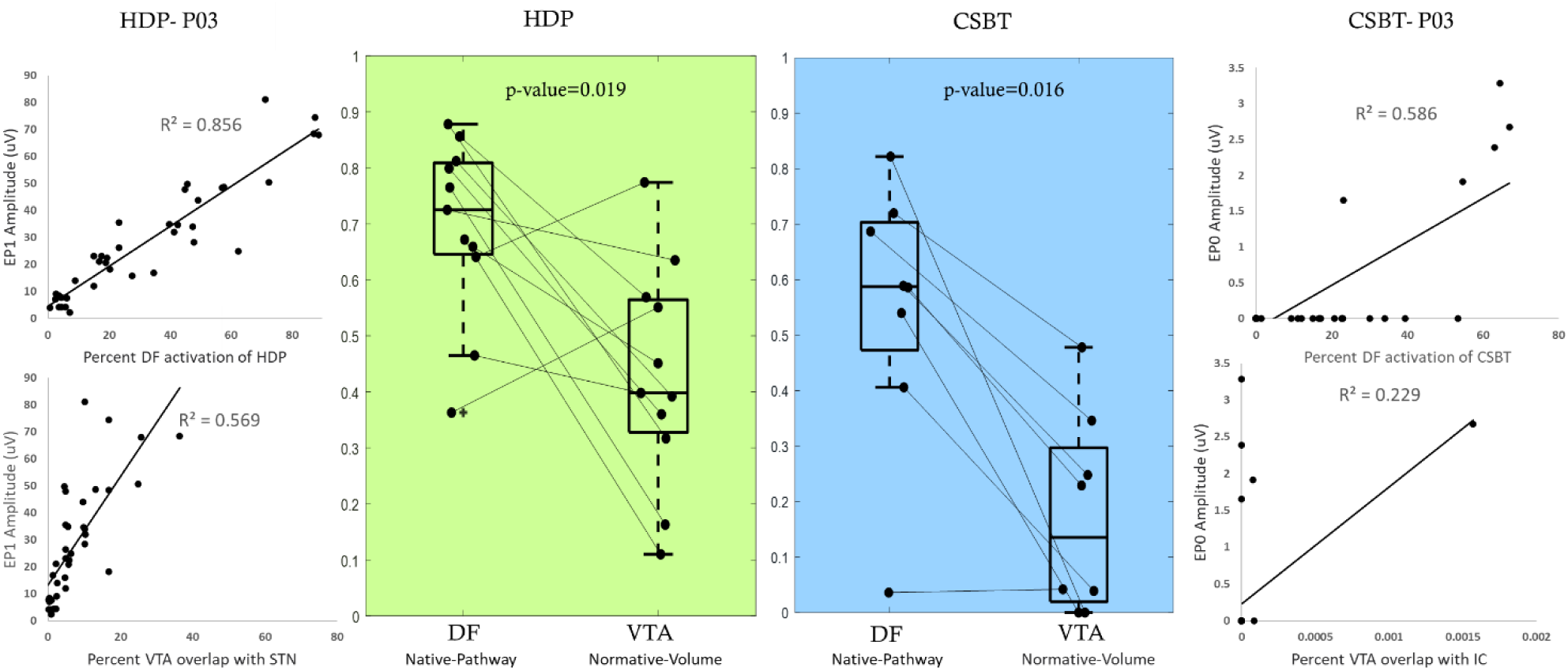
Comparison between the predictive performance of two common model types: DF-Native-Pathway and VTA-Normative-Volume for two pathways of interest: HDP (green-left) and CSBT (blue-right). Each data point in a box plot is the R2 value for one patient, i.e. the square of the correlation coefficient between the modeling activation predictions and experimental cEP amplitudes. Example correlation plots are shown for one patient (P03) comparing model predictions and cEP amplitudes for all stimulation settings.

We compared the models based on three factors to determine how each contributed to the observed performance differences (Fig. 7). To assess the importance of imaging space, we compared methodologies which were built in both native and normative spaces which resulted in two paired comparisons in each pathway: 1) DF-*Native*-Pathway vs DF-*Normative*-Pathway; 2) VTA-*Native*-Volume vs VTA-*Normative*-Volume. For HDP, the median *R*^2^ was 0.73 (IQR: 0.65-0.81) and 0.46 (IQR: 0.39-0.59) for DF-Pathway and VTA-Volume in native space, respectively, and both were significantly higher compared to 0.50 (IQR: 0.37-0.66) and 0.40 (IQR: 0.33-0.57) for their respective models in normative space. For CSBT, the *R*^2^ median was 0.59 (IQR: 0.47-0.70) and 0.31 (IQR: 0.06-0.38) for DF-Pathway and VTA-Volume in native space, respectively and both significantly higher compared to 0.41 (IQR: 0.32-0.51) and 0.14 (IQR: 0.02-0.30) for respective modeling methodology in normative space (Fig. 7-Top).

**Figure 7.**
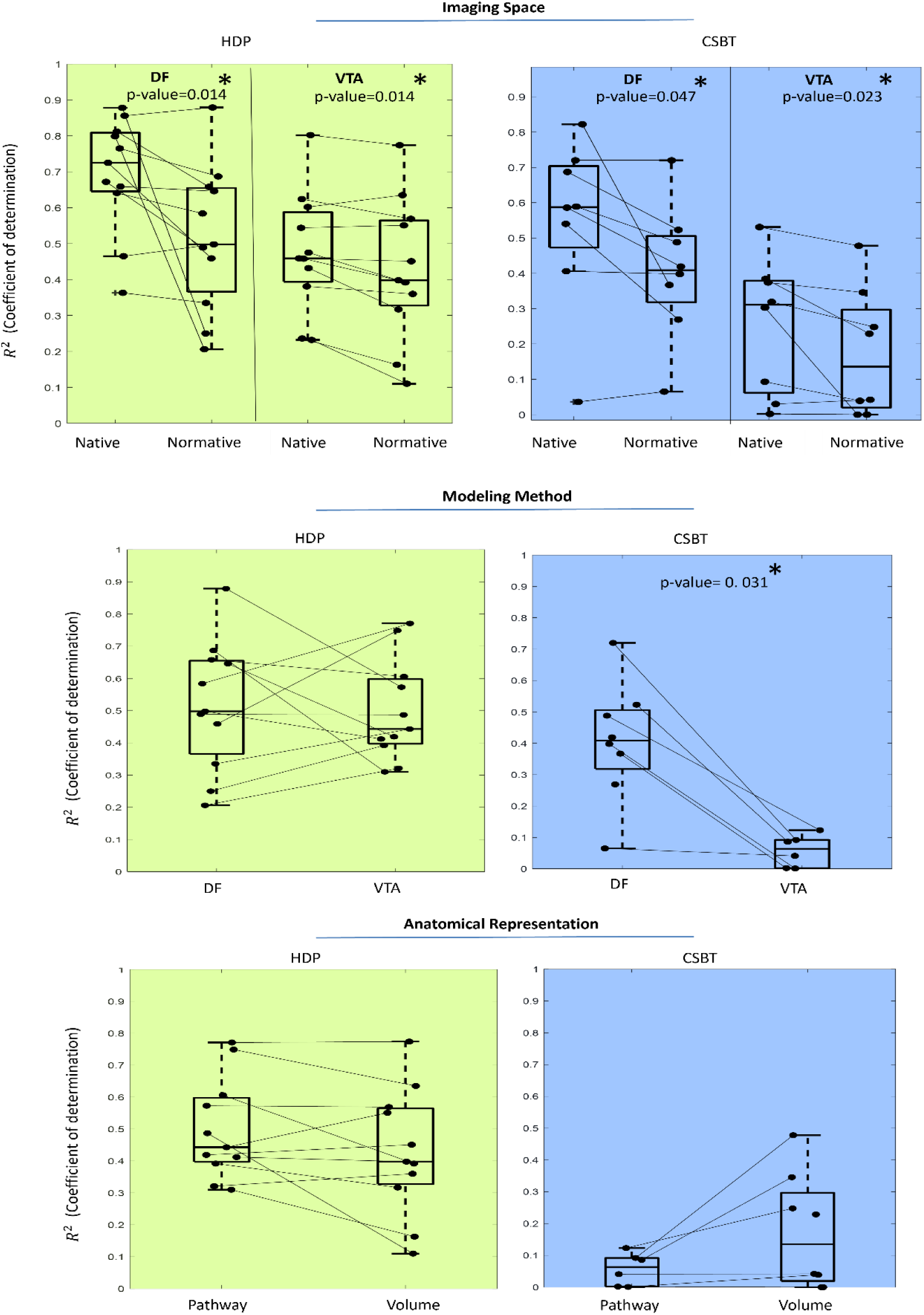
The effect of three key factors on model performance when predicting activation of HDP (green-left) and CSBT (blue-right) pathways. Significant differences are marked with asterisk (*). Top: Two model types – DF-Pathway and VTA-Volume were generated in native and normative space and in all cases the native model outperformed its normative counterpart. Middle: DF and VTA modeling methods were directly compared in normative space. DF outperformed VTAin predicting CSBT activation. Bottom: Pathway and volume anatomical representation were compared in VTA-Normative models with no significant differences.

To directly compare the effect of modeling method (DF vs VTA), the only possible pair of models was in the normative space with pathway representation. For HDP, the median *R*^2^ was 0.50 (IQR: 0.37-0.66) for DF and comparable to 0.44 (IQR: 0.40-0.60) for VTA model (Fig. 7-Middle/Right). For CSBT, the *R*^2^ median was 0.41 (IQR: 0.32-0.51) for DF which was significantly higher compared to 0.06 (IQR: 0.00-0.09) for VTA model (Fig. 7-Middle/Left). Also, the F-score metric showed significantly higher values for DF method with median of 0.52 (IQR: 0.34-0.72) compared to VTA at 0.10 (IQR: 0.00-0.28) (p-value = 0.008) (Fig. S1).

Finally, to evaluate the effect of anatomical representation (pathway vs structure volume) we used VTA models in normative space as the only possible pair for comparison among the implemented variations. For HDP, the median *R*^2^ was 0.44 (IQR: 0.40-0.60) for pathway model and comparable to 0.40 (IQR: 0.33-0.57) for volume model (Fig. 7. Bottom/Right). For CSBT, the *R*^2^median was 0.06 (IQR: 0.00-0.09) for pathway and comparable to 0.14 (IQR: 0.02-0.30) for volume model (Fig. 7-Bottom/Left). For CSBT, the F-score metric showed significantly higher values for the Volume model with median of 0.55 (IQR: 0.42-0.80) compared to Pathway model at 0.10 (IQR: 0.00-0.28) (p-value = 0.008) (Fig. S1).

## Discussion

We compared activation predictions from five different computational DBS model variations -differing in biophysical modeling method (DF vs VTA), imaging space (native vs normative), and anatomical representation (volume vs pathway) - with experimental measures of HDP and CSBT pathway activations from PD patients undergoing STN DBS surgery. In general, DF models were more accurate than VTA models, although there were some cases such as Normative-Pathway methodology for HDP activation for which the performance of VTA and DF were comparable. Furthermore, predictions were more accurate in the native compared to normative space, independent of the other methodologies. Overall, the DF models implemented in native space with pathway representation most accurately predicted experimental activations compared to all other methodologies, including the conventional VTA models implemented in normative space with volume representation.

Computational modeling using detailed multicompartment field-cable neural representations requires extensive computational time, resources, and specialized training and it is therefore difficult to execute for most practical applications (Patrick, 2024). As a result, simpler models have been proposed as a compromise to more effectively bridge computational approaches with clinical applications. The advent of open-source tools such as Lead-DBS (Horn et al., 2019; Neudorfer et al., 2023), SimNIBS (Saturnino et al., 2019), and ROAST (Nasimova & Huang, 2022), along with the increasing accessibility of electric field calculations using the finite-difference method (FDM) (Dimov et al., 2015) or finite element method (FEM), has helped streamline many aspects of brain stimulation modeling. VTA-based analyses are conceptually appealing due to their ability to provide a simple visual representation of “stimulation spread” (Åström et al., 2015a; Chaturvedi et al., 2013; Mädler & Coenen, 2012), leading to their widespread adoption in clinical research (Frankemolle, 2010) and clinical programming (Pourfar et al., 2015). However, our analysis shows that neglecting patient-specific DBS-induced electric fields and axonal trajectories in these models can significantly impair predictive performance. This oversight results in large biases that limit their accuracy for individualized analyses (Duffley et al., 2019; Gunalan et al., 2018a). Our results indicate that more recent “driving-force” (DF) models (Peterson et al., 2011; Warman et al., 1992), when implemented in native space with pathway representation, may better balance accuracy and simplicity, making them more suitable for clinical use (Howell, Choi, et al., 2019; Howell et al., 2021). The need for computational power that was limiting applicability of more realistic and complex models like DF in the early days is no longer an issue today. Furthermore, with advances in imaging techniques, models can now be built from high-resolution medical imaging (Coenen et al., 2021; Nordin et al., 2019; Wårdell et al., 2022), which allows models to be personalized. As the best predictive performance in our comparative analysis was achieved using the DF-Native-Pathway methodology, our subsequent analysis aimed to evaluate the effect of each dimension independently.

### Modeling in native space improves overall predictive performance

Independent of the modeling method or anatomical representation used, our analysis demonstrated that implementing methodologies in patient-specific (native) space resulted in better performance in predicting the experimental measures of HDP and CSBT activation compared to normative space. The superior performance of DBS modeling in patient-specific space highlights the importance of constructing individualized models rather than relying on generic, normalized atlases. Especially in VTA-based methods, the superiority of modeling in native space may be explained by the fact that while the non-linear transformation from patient space to normative space is applied to images and atlases (pathway or volumes), the electric field is computed without incorporating this transformation. This observation may also explain why recent efforts to use modeling software to correlate model predictions in normative space with PD motor symptom improvement and side effects for individual patients have been largely unsuccessful (Elias et al., 2021). It also aligns with the growing concern within the clinical community that employing DBS modeling based on aggregate patient data is inadequate for guiding surgical treatment in individual patients (Coenen et al., 2021; Elias et al., 2021; Rajamani et al., 2024).

### DF modeling has performance advantage for CSBT activation predictions

The performance advantage of the DF-models compared to VTA-models was more pronounced when predicting CSBT (Δr∼0.38) activation compared to HDP (Δr∼0.21). Additionally, in normative space the difference between DF and VTA models was statistically significant only for CSBT (Δr∼0.41). Several factors may have contributed to this result. First, VTA-based models can accurately estimate stimulation effects close to the electrode, but they underestimate activation of pathways or structures that are farther away (Gunalan et al., 2018b). This effect may be more pronounced by moving the analysis into the MNI normative space, which is an exceptionally large brain volume, resulting in CSBT streamlines residing even farther from the stimulating contact and resulting in fewer activated fibers. The VTA models underestimate the spread of stimulation effects whether using anatomical structure volume representation or a pathway atlas. This is likely because conceptually these models are similar -they calculate either how much structure or how many streamlines overlap with a fixed VTA. VTA models may underestimate stimulation effects at larger distance from the electrode because either voltage field estimate falls off too quickly or the voltage field threshold used to generate VTA is suboptimal (Gunalan et al., 2018b; Howell, Gunalan, et al., 2019). Specifically in the case of CSBT, a fundamental issue is that VTA algorithms are generic representations of stimulation spread, derived from a single fiber diameter in an isotropic brain medium. However, CSBT fibers are embedded in the highly anisotropic internal capsule, which alters the DBS voltage distribution (McIntyre et al., 2004) and substantially alters axonal response thresholds (Howell & McIntyre, 2016). Alternatively, DF models account for the fact that voltage distribution ‘seen’ by the axons varies along their trajectory. These factors help explain why reports using patient-specific DBS electric field models, explicitly representing brain anisotropy(Butson et al., 2007; Chaturvedi et al., 2010) reported a stronger association between CSBT activation and clinical side effect thresholds compared to simpler models (Béreau et al., 2020), which provided limited guidance in identifying capsular thresholds.

### Optimal DBS computational model may depend on specific application

Although we have demonstrated that using models with more realistic biophysical details in patient-specific space with pathway anatomical representation results in higher accuracy, this does not imply that other methodologies lack utility. The appropriate methodology depends on the specific application and the required level of accuracy. For example, the VTA-Volume method in patient-specific space showed reasonable performance (median r^2^ = 0.46) for HDP activation; however, for CSBT, its performance dropped significantly (median r^2^ = 0.31). Therefore, the selection of a methodology should consider, first, whether the focus is on therapeutic or side-effect pathways (or more generally near vs far pathways), and second, the degree of precision required for the specific clinical or research objective. Furthermore, in some cases using all patient-specific data is either not possible due to availability or quality issues or is hard to aggregate in a group-level analyses (Krauss et al., 2020). In these cases, some information should be restored from normative atlases and average brain templates. While studies attempting to correlate model-defined “sweet spots” with clinical outcomes have reported low correlation coefficients (Dembek et al., 2019; Ewert et al., 2018; Treu et al., 2020), they nonetheless achieved statistical significance. Additionally, sweet-spot targeting remains a common practice in clinical DBS, where volume-based methodologies have proven their utility (Pourfar et al., 2015; Waldthaler et al., 2021). Using VTA-based stimulation parameter selection has also been shown to diminish the cognitive and cognitive–motor declines associated with bilateral STN DBS (Frankemolle et al., 2010). However, it is noteworthy that such clinical improvement might also be mediated by correlating DBS lead locations rather than model-based activation estimates with best outcomes.

It is also noteworthy that when using the F1 score metric—focused on the presence of fiber activation rather than the degree of fiber activation —DF-Native-Pathway lost its superiority, with no significant difference between the methodologies (except when compared to VTA-Normative-Pathway, which performed worse than the others). This suggests that if the clinical goal is merely to predict the presence of side effects in a particular setting, most strategies can provide comparable predictive performance (assuming any internal capsule fiber activation results in a clinical side effect which is also unknown). However, if quantifying the degree of pathway activation is critical, DF-Native-Pathway remains the optimal choice based on our findings. Incorporating pathway-specific and connectivity-informed approaches can enable relating modeling outcomes to specific symptoms. For example, in PD, STN–SMA pathways relate to bradykinesia and rigidity improvement, while STN–motor cortex pathways mediate tremor relief (Akram et al., 2017). Or in essential tremor, Vim–motor cortex and Vim–cerebellum pathways are linked to tremor reduction (Al-Fatly et al., 2019). So, incorporating pathway-specific information and atlases, even with similar predictive performance, may reveal different aspects of the movement disorders compared to location- and volume-based approaches.

### Discrepancies between model predictions and clinical outcomes

While seeking the best computational model, it is important to reassess our expectations regarding the role of computational modeling in explaining or predicting clinical outcomes. When models poorly explain clinical outcomes, should we attribute the shortcomings solely to the way computational models are implemented, or do the limitations extend beyond modeling? Undoubtedly, models may be inaccurate, and with experimental evaluations such as those presented in this work, we can enhance their accuracy and standardize procedures for specific applications. However, even if neural activations and other brain phenomena can be modeled with high fidelity, model may still fall short in capturing sufficient anatomical or biophysical detail to fully explain underlying mechanisms. Furthermore, it is also possible that while the models are accurate in predicting neural activations, the questions we pose to them are inappropriate. Accurate modeling may be a necessary condition for predicting DBS clinical outcomes, but it is not a sufficient one. Additional information—such as the dynamic state of the broader basal ganglia network over time—might be essential for reliable outcome prediction. For instance, it remains unclear which specific neural pathway should be targeted to maximize therapeutic benefit. In STN DBS, potential targets include the subthalamo-pallidal efferent pathway (STN projection neurons), pallido-subthalamic afferents, the pallido-thalamic tract, and the motor cortico-subthalamic HDP. It is plausible that multiple pathways contribute to the overall therapeutic effect, or that certain pathways preferentially affect specific symptoms (Borgheai et al., 2025; Herrington et al., 2016). Quantifying therapeutic benefit presents additional challenges, as the severity of parkinsonian symptoms can fluctuate due to factors unrelated to the stimulation itself, such as medication levels, fatigue, and psychosocial factors. Moreover, while shifting from region-based to connectomic or pathway-based atlases has yielded promising results (Hines et al., 2024), further progress is required. For example, moving from pathway-based to network-level models that incorporate the topological (network) properties of the brain.

### Limitations

Our study has several limitations: 1) In implementing different models, we used two software packages and for consistency we used one structural and one pathway atlas. While these software programs capture the latest standards for implementing VTA and DF predictors, it is possible that our results are impacted by the specific tools we have chosen and, in the future, examining other alternatives could make the conclusions more generalizable. 2) Our statistical analysis was limited by relatively low number of patients which is a common limitation in intraoperative research. This especially may have affected results for CSBT pathway, where some activation measures were all zero either in-silico or in-vivo. 3) We focused on two specific pathways, while there are other clinically relevant pathways. A broader subthalamic pathway inclusion and activation measure could make the work more generalizable. 4) To keep DBS contact localization consistent across different models, we used localization tools in StimVision and then used the same coordinates for VTA modeling. While this localization matching helped compensate for discrepancies between the imaging pipelines used in DF- and VTA-based methods, it bypassed alternative localization and imaging strategies available in Lead-DBS, which could have influenced the results.

## Conclusion

While the DF-Native-Pathway model proved to be the most accurate method for quantitatively predicting experimental subcortical pathway activations, we believe that the choice of methodology should depend on the specific application and the required level of precision in clinical, surgical, or research settings. Our analysis showed that using normative brain space, instead of native (i.e., patient-specific) space, significantly diminished the accuracy of model predictions. Although the DF and VTA modeling methods exhibited comparable accuracy for the hyperdirect pathway, they diverged significantly in their predictions for the corticospinal tract, likely due to the VTA method’s limitations in estimating stimulation effects at greater distances from the electrode. These findings offer valuable guidance for developing more accurate models, facilitating reliable DBS outcome predictions, and advancing both clinical practice and scientific research.

## Data Availability

The data used in the present study will be made available upon publication of the manuscript in a peer-reviewed journal.

## Acknowledgement

We would like to thank Philip Starr, MD, PhD (UCSF) for sharing the intraoperative data, which contributed significantly to this work. This study was supported by NIH R01 NS125143 and NIH R37 NS116079.

## Supplementary Material

**Table S1.** Performance comparison of all the methodologies in predicting HDP activation.

| HDP | Native space |  | Normative space |  |  |
| --- | --- | --- | --- | --- | --- |
|  | DF -Pathway | VTA - Volume | DF - Pathway | VTA- -Volume | VTA-Pathway |
| P01 | 0.765 | 0.236 | 0.687 | 0.163 | 0.310 |
| P02 | 0.363 | 0.544 | 0.335 | 0.551 | 0.443 |
| P03 | 0.856 | 0.624 | 0.879 | 0.569 | 0.573 |
| P04 | 0.659 | 0.459 | 0.646 | 0.451 | 0.419 |
| P05 | 0.641 | 0.802 | 0.584 | 0.774 | 0.771 |
| P06 | 0.465 | 0.458 | 0.498 | 0.398 | 0.412 |
| P07 | 0.812 | 0.475 | 0.658 | 0.392 | 0.606 |
| P08 | 0.672 | 0.232 | 0.489 | 0.110 | 0.487 |
| P09 | 0.725 | 0.602 | 0.459 | 0.635 | 0.749 |
| P10 | 0.878 | 0.432 | 0.250 | 0.317 | 0.392 |
| P11 | 0.799 | 0.381 | 0.206 | 0.360 | 0.321 |
| Median | 0.725 | 0.459 | 0.498 | 0.398 | 0.443 |
| Q1 | 0.646 | 0.394 | 0.366 | 0.328 | 0.397 |
| Q3 | 0.809 | 0.588 | 0.655 | 0.565 | 0.598 |
| Mean | 0.694 | 0.477 | 0.517 | 0.429 | 0.499 |
| SD | 0.160 | 0.167 | 0.202 | 0.197 | 0.158 |

**Table S2.** Performance comparison of all the methodologies in predicting CSBT activation.

| CSBT | Native space |  | Normative space |  |  |
| --- | --- | --- | --- | --- | --- |
|  | DF -Pathway | VTA - Volume | DF - Pathway | VTA- -Volume | VTA-Pathway |
| P01 | 0.540 | 0.303 | 0.269 | 0.000 | NaN |
| P02 | 0.406 | 0.093 | 0.398 | 0.039 | 0.001 |
| P03 | 0.586 | 0.384 | 0.419 | 0.229 | NaN |
| P04 | 0.687 | 0.374 | 0.523 | 0.346 | 0.086 |
| P05 | 0.589 | 0.319 | 0.488 | 0.248 | 0.123 |
| P06 | 0.720 | 0.531 | 0.720 | 0.478 | 0.092 |
| P07 | NaN | NaN | NaN | NaN | NaN |
| P08 | 0.822 | 0.002 | 0.367 | 0.000 | 0.002 |
| P09 | 0.036 | 0.030 | 0.065 | 0.042 | 0.041 |
| P10 | NaN | NaN | NaN | NaN | NaN |
| P11 | NaN | NaN | NaN | NaN | NaN |
| Median | 0.588 | 0.311 | 0.408 | 0.136 | 0.064 |
| Q1 | 0.473 | 0.0615 | 0.318 | 0.0195 | 0.002 |
| Q3 | 0.7035 | 0.379 | 0.5055 | 0.297 | 0.092 |
| Mean | 0.548 | 0.255 | 0.406 | 0.173 | 0.058 |
| SD | 0.242 | 0.191 | 0.191 | 0.180 | 0.051 |

**Figure S1.**
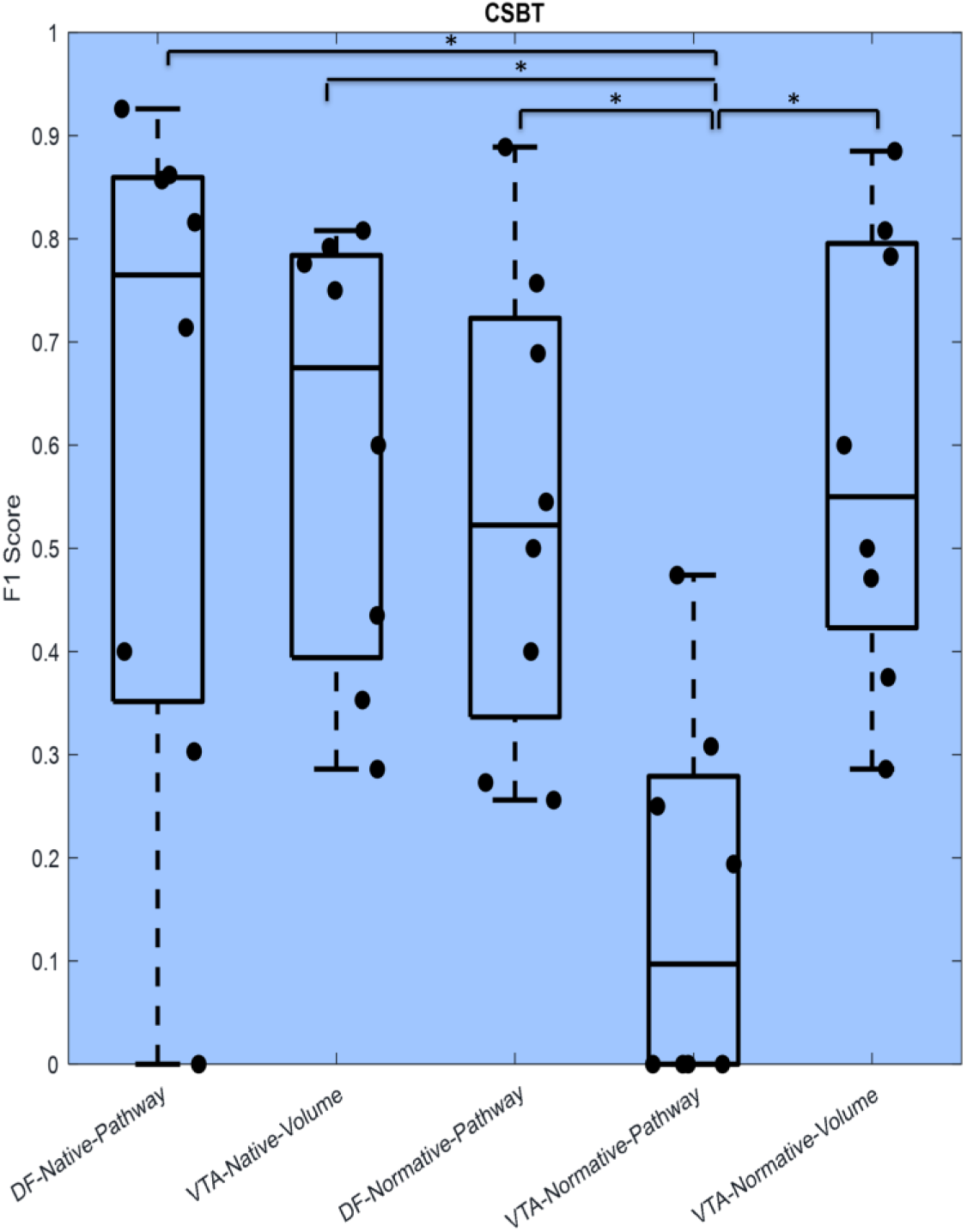
F1 Score comparison between the performances of all methodologies in CSBT. The pairs with significant differences are shown with bracket and asterisk (*).

